# Prevalence and drug-resistance patterns of methicillin-resistant *Staphylococcus aureus* and extended-spectrum β-lactamase producing *Enterobacteriaceae* among raw vegetables

**DOI:** 10.1101/2025.01.28.25321130

**Authors:** Duressa Shafi Ahmed, Alemitu Beyene Gebre, Hadbacho Tsegaye Erancho, Habtamu Teshale Shinato, Moges Desta Ormago

## Abstract

**Background:** Antimicrobial resistance is a top global public health and development threat. Antibiotic-resistant bacteria in food items, such as vegetables, have become a public health issue. Contamination of vegetables sold in various places can cause foodborne illnesses in many countries, including Ethiopia, and affect the health of consumers. Many bacterial pathogens are now resistant to multiple drugs. In recent years, several multi-drug resistant bacterial pathogens with epidemiological significance have emerged, such as MRSA and ESBL-PE.

**Methods:** A cross-sectional study was conducted in Aroge Gebeya Market, Hawassa City, from March to May 2024. Six types of raw vegetables with a total of 216 (6×36) were collected. Identification of methicillin-resistant *Staphylococcus aureus* and extended-spectrum β-lactamase producing *Enterobacteriaceae* and drug resistance patterns were done by using the Kirby-Bauer disk diffusion method. Data on sociodemographic variables and hygiene practices were collected using semi-structured questionnaires on Kobo Toolbox Software and imported to SPSS Software Version 29.0.1 for analysis.

**Results:** The overall prevalence of MRSA and/or ESBL-PE among vegetables was 18.1% (39/216). Among the isolates, 56.9% (91/160) were MDR, 29.2% (14/48) were MRSA, and 24.1% (27/112) were ESBL producers. The most frequently detected isolates were *S. aureus* 30.0% (48/160), *Enterobacter spp.* 15.6% (25/160), *Citrobacter spp.* 15.0% (24/160), *E. coli* 10.6% (17/160), *Salmonella spp.* 10.0% (16/160) and *Klebsiella spp.* 8.1% (13/160). The highest frequency of bacterial isolates was found in tomatoes 21.9% (35/160), kale 18.1% (29/160), and lettuce 16.9% (27/160). Vegetables bought from vendors with untrimmed nails were significantly more likely to have a higher prevalence of MRSA and/or ESBL-PE compared to those from vendors with trimmed nails (AOR = 4.123; 95% CI: 1.681 - 10.109, p = 0.002).

**Conclusion:** This study found that vegetables were notable for the presence of MDR bacteria, specifically ESBL-PE and MRSA. Consequently, we strongly recommend that local health authorities develop and implement effective control strategies to mitigate these resistant bacteria among vegetables.

## Background

Antimicrobial resistance (AMR) is one of the top global public health and development threats that happens when microorganisms, such as bacteria, viruses, fungi and parasites, evolve mechanisms that protect them from the effects of antimicrobials and no longer respond to antimicrobial medicines (1). AMR has now emerged as a chronic public health problem globally, with the forecast of 10 million deaths per year globally by 2050 (2). The problem is not restricted to human-linked habitats, since other ecosystems, including animals, soil and water bodies contribute to the origin, spread and maintenance of AMR (3,4). Antibiotic resistance, which is the crucial aspect of AMR, has been attributed to the misuse and overuse of antibiotics which puts selective pressure on bacterial pathogens leading to the emergence of resistance (1,5,6). The recent emergence of antibiotic-resistant bacteria in food items, including vegetables, has evolved into a public health concern. (7).

Vegetables are favored for their ease of use and wide acceptance; however, their lack of a terminal heating process before consumption makes them prone to high levels of microbial contamination and cross-contamination, potentially leading to food-borne pathogen outbreaks (8). These issues are further compounded by inadequate handling practices and the absence of stringent food safety regulations in these environments, exacerbating the risk of widespread contamination. The rise in food-borne illnesses linked to vegetables in many countries, including Ethiopia, is due to contamination that can occur in all vegetables sold in public markets, supermarkets, and roadside stalls, leading to significant health issues for consumers (9).

Multi-drug resistance (MDR), the phenomenon where microbes become resistant to several drugs, is now common among many bacterial pathogens (10). In the last few decades, several epidemiologically significant MDR bacterial pathogens have emerged. These include methicillin-resistant Staphylococcus aureus (MRSA) and extended-spectrum β-lactamase producing *Enterobacteriaceae* (ESBL-PE) (10,11).

*Staphylococcus aureus* species are non-spore-forming Gram-positive bacteria in the form of cocci; they are non-mobile, mesophilic, biofilm-forming, and facultative anaerobes that produce enterotoxins (12). *S. aureus* can be transmitted by contaminated food, and its transmission is mainly due to the poor handling of food during processing (13). It can release a variety of heat-stable staphylococcal enterotoxins (SEs) into food (14,15). The consumption of food contaminated with these toxins produced by *S. aureus* can lead to staphylococcal food poisoning, which may cause severe gastroenteritis, nausea, vomiting, diarrhea, and abdominal pain within 1 to 6 h after the consumption of contaminated food (13,16), and it was recognized as one of the most prominent culprits in food poisoning outbreaks worldwide (17). *S. aureus* can also lead to other diseases; some of them are severe, such as sepsis, endocarditis and necrotizing pneumonia (13,18).

Currently, this microorganism’s ability to develop resistance to antibiotics is notorious (19). The resistance is usually acquired by horizontal gene transfer, although mutation and selection are also important (20). Infections caused by resistant strains are common in epidemic waves by one or more clones; methicillin-resistant *Staphylococcus aureus* (MRSA) is prominent in epidemic waves, being historically associated with hospitals and health units (healthcare-associated MRSA (HA-MRSA)) (13,20). Nowadays, it has emerged as a cause of community-acquired infections (CA-MRSA), spreading rapidly among healthy individuals and its presence is a cause of concern due to resistance to various antibiotics, limiting treatment (13).

Methicillin-resistant *S. aureus* (MRSA) strains were first described in 1961 (21–23), and they were characterized by an altered penicillin-binding protein (PBP2a), which had reduced affinity for methicillin and, thus, could continue peptidoglycan synthesis uninterrupted in the presence of this drug (21–24). MRSA can develop through the acquisition of determinants by horizontal gene transfer of mobile genetic elements (MGEs); from transposons and the staphylococcal cassette chromosome element (25,26); from chromosomal mutations that alter drug binding sites on molecular targets; or by increasing expression of endogenous efflux pumps (25,27).

Another epidemiologically significant MDR bacterial pathogen is ESBL-PE (10,28). This pathogen has emerged as a serious problem not only in nosocomial infections but also in community settings and obtained intensive attention in recent years (8). Raw vegetables can be contaminated by ESBL-producing *Enterobacteriaceae* through various sources, such as contaminated biological fertilizer or water, poor hygiene practices of the workers involved in food production, ineffective food safety management and unsanitary vegetable packaging or display (29).

β-Lactamases are bacterial enzymes that inactivate β-lactam antibiotics by hydrolysis, rendering them ineffective (30). The primary factor contributing to β-lactam resistance in Gram-negative bacteria is β-lactamase production, which evolves due to selection pressure from various β-lactam agents (31). β-Lactamases differ in their substrate profiles, inhibitor profiles, and sequence homology (30), which has led to the classification of β-lactamases into Ambler classes A, B, C, and D based on amino acid sequence homology, and Bush-Jacoby-Medeiros groups 1, 2, and 3 based on substrate and inhibitor profiles. Most ESBLs belong to Ambler class A, including the SHV and TEM types (31). ESBLs confer resistance to oxyimino-cephalosporins and monobactams but not to cephamycin or carbapenems, and they can be inhibited by classical β-lactamase inhibitors such as clavulanic acid, sulbactam, and tazobactam (31–34).

Despite the known risks, there is a scarcity of research in low-income countries evaluating foodborne pathogens and their antibiotic resistance patterns. The burden of MRSA and ESBL-producing Enterobacteriaceae in vegetables in Ethiopia is underexplored. To address these gaps, this study aimed to present fundamental statistics on the prevalence of MRSA and ESBL-PE in raw vegetables commonly sold at the Aroge Gebeya market in Hawassa, Ethiopia. By providing accurate and up-to-date data on these multidrug-resistant pathogens, the study seeks to inform and support health planners and caregivers in developing evidence-based interventions to combat AMR. Such data are critical for raising awareness among health professionals and communities about the potential risks associated with consuming contaminated vegetables and for contributing to future large-scale AMR studies.

## Methods

### Study design and study area

A cross-sectional study was conducted from March to May 2024. The study was conducted in Aroge Gebeya, a market in Hawassa, the capital city of Sidama regional state, which is located approximately 273 km (170 mi) south of Addis Ababa. According to the Central Statistics Agency, the city population estimate in 2023 was 577,075 (38). Hawassa City boasts a rich tapestry of traditional markets, with Aroge Gebeya emerging as a prominent hub. This market serves as the primary food supplier for the city’s residents, making it an ideal location for research. Its extensive size, accessibility, and economic significance contribute to its suitability. The market’s unique blend of permanent and temporary stalls, coupled with its daily operation and high foot traffic, offers a dynamic environment for observation.

### Data and sample collection

Data on socio-demographic characteristics and sanitary risk variables were collected using semi-structured questionnaires on Kobo Toolbox software (Version 2.022.44g). A total of 216 vegetables were bought from Aroge Gebeya market: 36 each of tomatoes, potatoes, carrots, red onions, kale, and lettuce. For carrots, red onions, tomatoes, and potatoes, half a kilogram of each was purchased, while kale and lettuce were bought in bunches. The marketplace was divided into distinct segments, and samples were selected from each segment using a stratified random sampling technique with a computerized random number generator. During the data collection process, vendors were instructed to randomly select vegetables as they usually sell them to customers. Vendors were actively monitored to prevent them from taking vegetables only from one side or choosing exclusively fresher or older samples. The vegetables were placed in clean plastic bags with labels and then transported in a cold box at low temperatures to the Microbiology Laboratory of Hawassa University College of Medicine and Health Sciences (HUCMHS).

### Laboratory testing method

#### Sample preparation

To prepare each vegetable sample for analysis, the initial step was aseptically opening the packaging bags using a sterile stainless-steel knife. One sample from every half kilogram of packed vegetables (tomato, potato, carrot, and red onion) or bunches (kale and lettuce) of vegetables was then randomly selected using a computer random number generator. Next, the selected vegetables were sliced or cut into small pieces using a sterilized tray and knife while wearing latex gloves for sanitary purposes. Subsequently, 25 g of each vegetable sample was weighed and enriched in 225 mL of buffered peptone water (BPW; Oxoid Ltd., Basingstoke, Hampshire, U.K.). The mixture was homogenized using a stomacher device (Seward Ltd., U.K.) and then incubated at 37°C for 24 hours. This process followed the guidelines outlined in the Bacteriological Analytical Manual (BAM), approved and accepted by the FDA, ensuring accurate identification of bacterial isolates (40).

#### Isolation and identification of bacteria

Following homogenization and incubation, a loopful (approximately 10 microliters) of each sample was inoculated onto solidified MacConkey Agar (Oxoid Ltd., Basingstoke, Hampshire, UK) and Mannitol Salt Agar (Central Drug House (P) Ltd.). Pure colonies were obtained and key characteristics were recorded. *Enterobacteriaceae* were identified based on colonial morphology and pigmentation, carbohydrate fermentation, H2S production, citrate utilization, motility, indole formation, lysine decarboxylase, and urea hydrolysis. *Staphylococcus aureus* was differentiated by colonial characteristics, as well as catalase, coagulase, and oxidase tests (40).

#### Antimicrobial susceptibility testing

Antimicrobial susceptibility testing (AST) was carried out by using the Kirby-Bauer disc diffusion method as recommended by Clinical and Laboratory Standards Institute (CLSI) guidelines (41). Pure culture colonies of 24-hour growth were suspended in a tube with 4 mL of physiological saline to get bacterial inoculums equivalent to 0.5 McFarland turbidity standards. A sterile cotton swab was dipped and rotated across the wall of the tube to avoid excess fluid, and evenly inoculated on Muller-Hinton agar (Biomark Laboratories), and then the antibiotic disks were placed on MHA plates.

The antibiotics disks (Oxoid Ltd., Basingstoke, Hampshire, UK), were used for both *S. aureus* isolates and *Enterobacteriaceae* isolates were based on the CLSI guidelines and their wide prescription in Ethiopia. Accordingly, ciprofloxacin (CIP, 5µg), trimethoprim-sulfamethoxazole (SXT, 25μg), gentamicin (CN, 10µg), clindamycin (CC, 2µg), cefoxitin (FOX, 30µg) and erythromycin (ERY, 15μg) were used for *S. aureus* isolates. On the other hand, trimethoprim-sulfamethoxazole (SXT, 25μg), ciprofloxacin (CIP, 5µg), meropenem (MER, 10μg), tetracycline (TET, 30μg), ceftriaxone (CRO, 30µg), cefotaxime (CTX, 30µg) and ceftazidime (CAZ, 30µg) were used for *Enterobacteriaceae* isolates. The plates were then incubated at 37°C for 24 hours. Diameters of the zone of inhibition around the disks were measured using a ruler. The interpretation of the results of the AST was based on the standardized table supplied by the national committee for CLSI criteria as sensitive, intermediate, and resistant (41).

#### Screening for Methicillin-Resistant *Staphylococcus aureus* (MRSA)

All isolates of S. aureus were screened for methicillin resistance using cefoxitin discs (30μg) with the Kirby-Bauer disk diffusion technique. Isolates with a zone of inhibition ≤21mm around the cefoxitin disc were identified as methicillin-resistant (41).

#### Inducible Clindamycin Resistance (ICR)

Inducible clindamycin resistance (ICR) was identified using a D-test, which involves the application of erythromycin (ERY, 15μg) and clindamycin (CC, 2μg) discs spaced 15−26 mm apart on Muller-Hinton Agar (MHA), as per CLSI guidelines. Flattening of the zone of inhibition adjacent to the erythromycin disc and cloudy growth within the zone of inhibition around clindamycin indicate ICR (41).

#### Screening of ESBL-Producing *Enterobacteriaceae*

Each *Enterobacteriaceae* isolate with reduced susceptibility to cefotaxime and/or ceftazidime was included as a potential ESBL producer. Isolates with an inhibition zone size of ≤22 mm for ceftazidime (30μg) and/or ≤27 mm for cefotaxime (30μg) were considered potential ESBL producers. To screen for ESBL production, discs of ceftazidime (30μg) and cefotaxime (30μg) were placed 25 mm apart on Muller-Hinton agar plates prepared with a bacterial suspension adjusted to 0.5 McFarland turbidity standards (41).

#### Confirmation of ESBL Production

In addition to the individual antibiotic discs, combination discs containing ceftazidime/clavulanic acid (30/10μg) and cefotaxime/clavulanic acid (30/10μg) were placed on the agar plates. The plates were incubated overnight at 37°C for 18-24 hours. Bacterial isolates were confirmed as ESBL producers if there was an increased zone of inhibition diameter of ≥5 mm for the combination discs compared to ceftazidime or cefotaxime alone, following CLSI guidelines (41).

### Data Analysis and Interpretation

Data from the Kobo Toolbox were imported into SPSS Statistics for Windows, Version29.0.1 (IBM Corp., Armonk, NY). The frequency distribution and the prevalence were calculated. Different statistical tables were used to display quantitative values. The socio-demographic and sanitary risk variables for the overall prevalence of MRSA and/or ESBL-PE among vegetables were assessed using a bivariate logistic regression analysis. The factors with a P-value of ≤0.25 were further investigated using multivariable logistic regression, as this threshold allows for the inclusion of potentially relevant variables that might not meet stricter significance levels, thus ensuring that important factors are not prematurely excluded from further analysis. Adjusted odds ratio (AOR), 95% CI, and P-value ≤0.05 were used to identify the significant association.

### Operational definitions

**Multi-drug resistance (MDR)**: is the resistance to at least one antibiotic in three or more different antimicrobial groups by microorganisms, with MRSA and ESBL-PE being typical examples (1).

**Overall prevalence of MRSA and/or ESBL-PE among vegetables:** refers to the proportion of vegetables that have only MRSA, only ESBL-PE, or both MRSA and ESBL-PE.

**Intact vegetables**: are those that haven’t been cut, sliced, or otherwise altered from their original form, retaining their natural outer layer.

**Not intact vegetables**: are vegetables that have been cut, sliced, or in any way processed.

**Vegetables that were spoiled and sorted separately**: are vegetables that have gone bad and are sorted separately, typically showing signs like discoloration, off-odors, sliminess, or mold growth.

## Results

### Socio-demographic characteristics of vendors and conditions of vegetables

In this study, among 216 vendors, 165 (76.4%) were females and the age of the vendors ranged from 15 to 60 years, with a mean age of 31.4 years and standard deviation (SD) of 9.8 years. Most of the vendors do not wash their vegetables before display 156 (72.2%). The majority of vendors display their vegetables without stall 133 (61.6%) (Table 1).

**Table 1.**
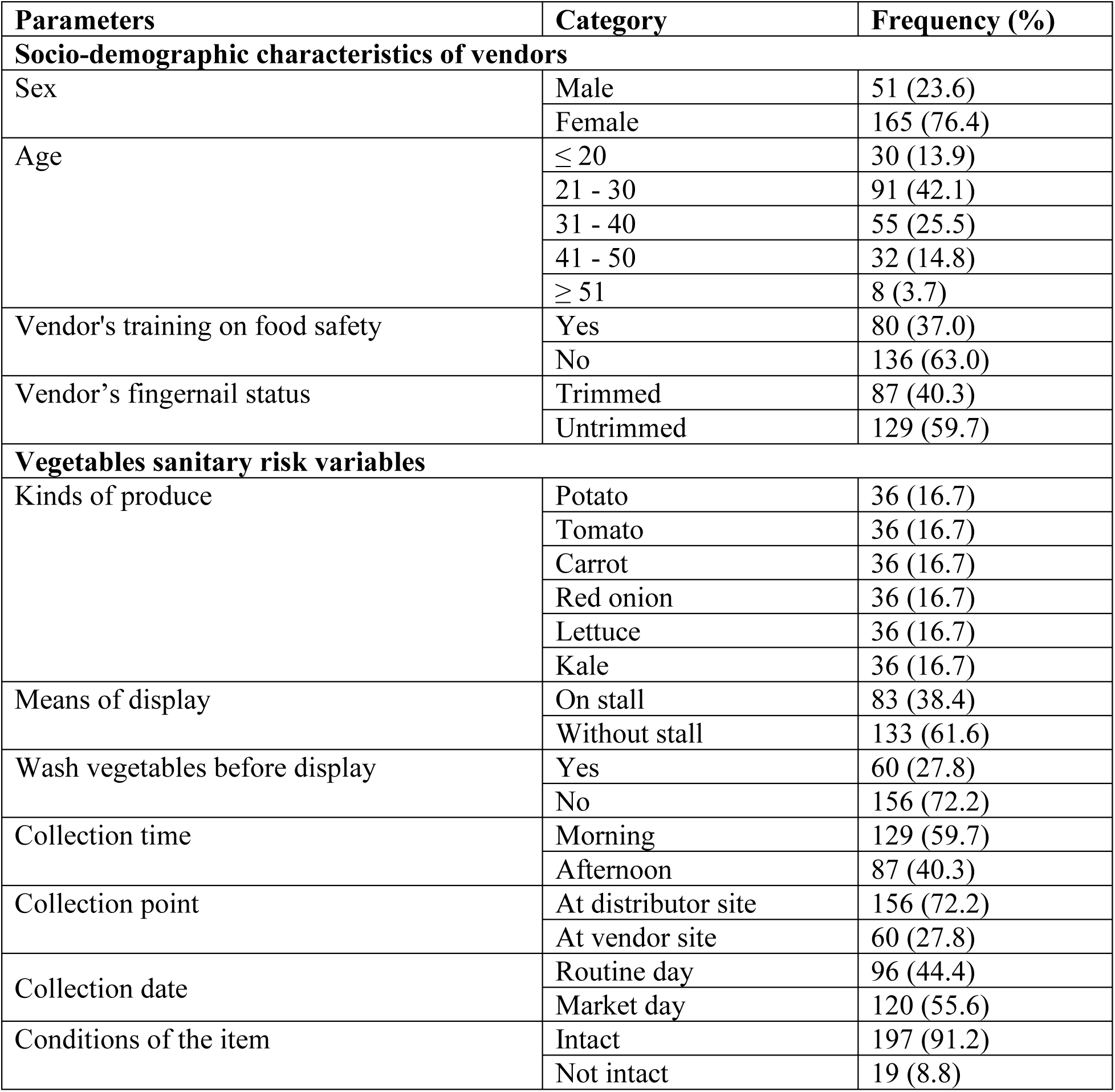
Socio-demographic characteristics of vendors and vegetables sanitary risk variables in Aroge Gebeya, Hawassa, Ethiopia, 2024.

| Parameters | Category | Frequency (%) |
| --- | --- | --- |
| **Socio-demographic characteristics of vendors** |  |  |
| Sex | Male | 51 (23.6) |
|  | Female | 165 (76.4) |
| Age | ≤ 20 | 30 (13.9) |
|  | 21 - 30 | 91 (42.1) |
|  | 31 - 40 | 55 (25.5) |
|  | 41 - 50 | 32 (14.8) |
|  | ≥ 51 | 8 (3.7) |
| Vendor's training on food safety | Yes | 80 (37.0) |
|  | No | 136 (63.0) |
| Vendor's fingernail status | Trimmed | 87 (40.3) |
|  | Untrimmed | 129 (59.7) |
| **Vegetables sanitary risk variables** |  |  |
| Kinds of produce | Potato | 36 (16.7) |
|  | Tomato | 36 (16.7) |
|  | Carrot | 36 (16.7) |
|  | Red onion | 36 (16.7) |
|  | Lettuce | 36 (16.7) |
|  | Kale | 36 (16.7) |
| Means of display | On stall | 83 (38.4) |
|  | Without stall | 133 (61.6) |
| Wash vegetables before display | Yes | 60 (27.8) |
|  | No | 156 (72.2) |
| Collection time | Morning | 129 (59.7) |
|  | Afternoon | 87 (40.3) |
| Collection point | At distributor site | 156 (72.2) |
|  | At vendor site | 60 (27.8) |
| Collection date | Routine day | 96 (44.4) |
|  | Market day | 120 (55.6) |
| Conditions of the item | Intact | 197 (91.2) |
|  | Not intact | 19 (8.8) |

### Frequency and distribution of bacterial isolates among vegetables

Among the 160 bacterial isolates, the most frequently detected were *S. aureus* 30.0% (48/160), *Enterobacter spp.* 15.6% (25/160), *Citrobacter spp.* 15.0% (24/160*), Escherichia coli* 10.6% (17/160), *Salmonella spp.* 10.0% (16/160), and *Klebsiella spp.* 8.1% (13/160). The highest frequency of bacterial isolates was found in tomatoes 21.9% (35/160), kale 18.1% (29/160), and lettuce 16.9% (27/160). *S. aureus* was the most prevalent isolate in tomatoes 22.9% (11/48), kale 20.8% (10/48), and red onions 18.8% (9/48). Among the *Enterobacteriaceae* isolates, *E. coli* was most frequently detected in kale 47.1% (8/17). *Enterobacter spp.* were predominantly isolated from potatoes 28.0% (7/25) and carrots 20.0% (5/25). *Citrobacter spp.* were mainly found in tomatoes 25.0% (6/24) and carrots 20.8% (5/24) (Table 2).

**Table 2.** Distribution of bacterial isolates among vegetables in Aroge Gebeya, Hawassa, Ethiopia, 2024. The values are provided as numbers (n) and percentages (%).

| Isolates (n) | Potato | Tomato | Carrot | Red onion | Lettuce | Kale | Total |
| --- | --- | --- | --- | --- | --- | --- | --- |
| *S. aureus* | 8 (16.7) | 11 (22.9) | 4 (8.3) | 9 (18.8) | 6 (12.5) | 10 (20.8) | 48(30.0) |
| *Enterobacter spp.* | 7 (28.0) | 4 (16.0) | 5 (20.0) | 4 (16.0) | 4 (16.0) | 1 (4.0) | 25(15.6) |
| *Citrobacter spp.* | 3 (12.5) | 6 (25.0) | 5 (20.8) | 1 (4.2) | 4 (16.7) | 5 (20.8) | 24(15.0) |
| *Escherichia coli* | 0 (0.0) | 3 (17.6) | 0 (0.0) | 2 (11.8) | 4 (23.5) | 8 (47.1) | 17(10.6) |
| *Salmonella spp.* | 1 (6.3) | 4 (25.0) | 3 (18.8) | 4 (25.0) | 3 (18.8) | 1 (6.3) | 16(10.0) |
| *Klebsiella spp.* | 1 (7.7) | 2 (15.4) | 3 (23.1) | 1 (7.7) | 3 (23.1) | 3 (23.1) | 13(8.1) |
| *Providencia spp.* | 2 (22.2) | 2 (22.2) | 2 (22.2) | 2 (22.2) | 1 (11.1) | 0 (0.0) | 9(5.6) |
| *Proteus spp.* | 2 (25.0) | 3 (37.5) | 0 (0.0) | 0 (0.0) | 2 (25.0) | 1 (12.5) | 8(5.0) |
| Total | 24(15.0) | 35(21.9) | 22(13.8) | 23(14.4) | 27(16.9) | 29(18.1) | 160(100.0) |

### Prevalence of MDR, MRSA and ESBL-PE

The overall prevalence of MRSA and/or ESBL-PE among vegetables was found to be 18.1% (39/216). Only two vegetable samples (one red onion and one lettuce) contained both MRSA and ESBL-PE simultaneously; the remainder contained either MRSA, ESBL-PE, or neither. Of the 160 bacterial isolates, 56.9% (91/160) were MDR, 29.2% (14/48) were MRSA, 98.2% (110/112) were screened for ESBL production, and 24.1% (27/112) were confirmed as ESBL producers. Among the isolates, *Salmonella spp.* (81.3%; 13/16), *Citrobacter spp.* (79.2%; 19/24), and Klebsiella spp. (69.2%; 9/13) showed the highest rates of MDR. Klebsiella spp. (46.2%; 6/13), *E. coli* (35.2%; 6/17), and *Salmonella spp*. (25.0%; 4/16) were among the isolates showing the highest rates of confirmed ESBL production (Table 3). The highest number of confirmed ESBL producers was found in tomatoes (22.2%; 8/36), followed by red onions (13.9%; 5/36). Additionally, the highest number of MRSA isolates was found in tomatoes (11.1%; 4/36) and red onions (11.1%; 4/36) (Table 4).

**Table 3.** Prevalence of MDR, MRSA and ESBL-PE among vegetables in Aroge Gebeya, Hawassa, Ethiopia, 2024. The values are provided as numbers (n) and percentages (%).

| Isolate(n) | MDR | MRSA | Screened for ESBL | Confirmed ESBL |
| --- | --- | --- | --- | --- |
| *S. aureus* (48) | 14 (29.2) | 14 (29.2) | NA | NA |
| *Enterobacter* spp. (25) | 16 (64.0) | NA | 24 (96.0) | 4 (16.0) |
| *Citrobacter* spp. (24) | 19 (79.2) | NA | 24 (100.0) | 4 (16.7) |
| *Escherichia coli* (17) | 10 (58.8) | NA | 16 (94.1) | 6 (35.3) |
| *Salmonella* spp. (16) | 13 (81.3) | NA | 14 (87.5) | 4 (25.0) |
| *Klebsiella* spp. (13) | 9 (69.2) | NA | 13 (100.0) | 6 (46.2) |
| *Providencia* spp. (9) | 6 (66.7) | NA | 9 (100.0) | 2 (22.2) |
| *Proteus* spp. (8) | 4 (50.0) | NA | 8 (100.0) | 1 (12.5) |
| Total (160) | 91 (56.9) | 14 (29.2) | 108 (96.4) | 27 (24.1) |

**Note:** MDR: Multi-drug Resistance, MRSA: Methicillin Resistant *S. aureus*, ESBL-PE: Extended Spectrum β-lactamase Producing *Enterobacteriaceae*, NA: Not Applicable.

**Table 4.** Distribution of MRSA and ESBL-PE among vegetables in Aroge Gebeya, Hawassa, Ethiopia, 2024. Values are provided as numbers (n) and percentage (%).

| Types of vegetables | ESBL confirmed | MRSA |
| --- | --- | --- |
| Tomato (36) | 8 (22.2) | 4 (11.1) |
| Potato (36) | 3 (8.3) | 2 (5.6) |
| Carrot (36) | 3 (8.3) | 1 (2.8) |
| Red onion (36) | 5 (13.9) | 4 (11.1) |
| Kale (36) | 4 (11.1) | 2 (5.6) |
| Lettuce (36) | 4 (11.1) | 1 (2.8) |
| Total (216) | 27 (12.5) | 14 (6.5) |

**Note:** MRSA: Methicillin Resistant *S. aureus*, ESBL-PE: Extended Spectrum β-lactamase Producing *Enterobacteriaceae*.

### Antimicrobial resistance of bacterial isolates

The highest level of resistance was observed by *Enterobacteriaceae* with cefotaxime (50.0%; 56/112), ceftazidime (46.4%; 52/112) and ceftriaxone (34.8%; 39/112). *S. aureus* showed the highest resistance to cefoxitin at (29.2%; 14/48) while showing no resistance to ciprofloxacin, trimethoprim-sulfamethoxazole and gentamycin. However*, Enterobacteriaceae* isolates showed lower resistance to ciprofloxacin (14.2%; 16/112) and trimethoprim-sulfamethoxazole (21.4%; 24/112) (Table 5). Of the bacteria isolates, (5.0%; 8/160) were resistant to five or more antibiotics, while (30.6%; 49/160) isolates were not resistant to any of the antibiotics tested. There were also resistant bacteria for one (23.8%; 38/160), two (15.6%; 25/160), three (13.8%; 22/160) and four (11.3%; 18/160) antibiotics (Table 6). Out of the total 14 MRSA isolates, 14 (100%), 12 (85.7%), and 10 (71.4%) were susceptible to gentamycin, trimethoprim-sulfamethoxazole, and ciprofloxacin, respectively. Conversely, 5 (35.7%) and 2 (14.3%) of MRSA isolates were resistant to clindamycin and erythromycin, respectively (Figure 1). Among the total MRSA isolates, 2 out of 14 (14.3%) showed inducible clindamycin resistance (ICR), whereas none of the methicillin-susceptible *S. aureus* (MSSA) isolates showed ICR (Table 7). Of the 27 ESBL-PE isolates, 19 (70.4%) and 13 (48.1%) were susceptible to trimethoprim-sulfamethoxazole and ceftriaxone, respectively. On the other hand, 20 (74.1%), 16 (59.3%), and 11 (40.7%) of ESBL-PE isolates were resistant to cefotaxime, ceftazidime, and ceftriaxone, respectively (Figure 2).

**Figure 1.**
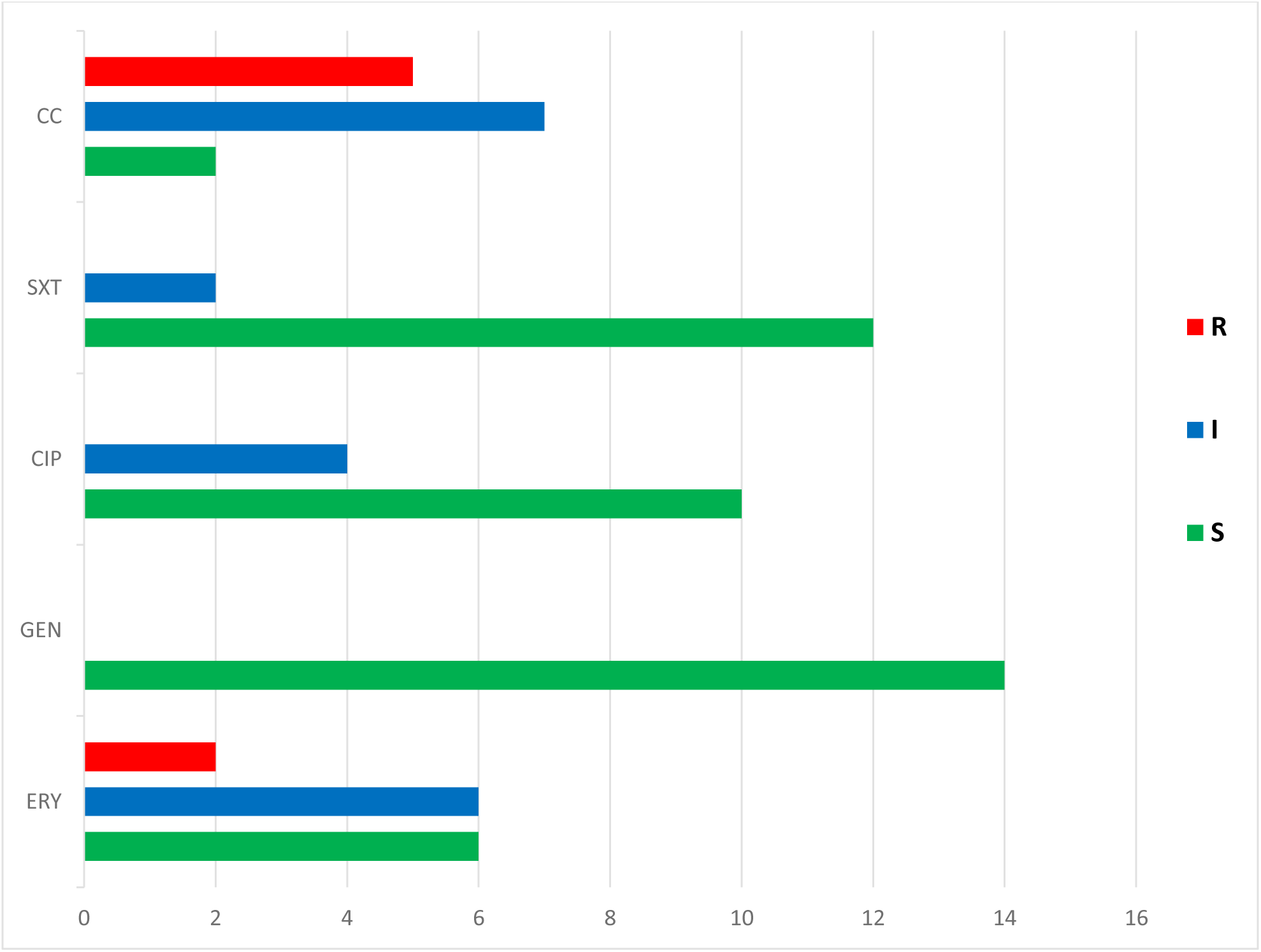
Antimicrobial susceptibility patterns of MRSA isolated from vegetables sold in Aroge Gebeya, Hawassa, Ethiopia, 2024. **Note:** R: Resistance, I: Intermediate, S: Susceptible

**Figure 2.**
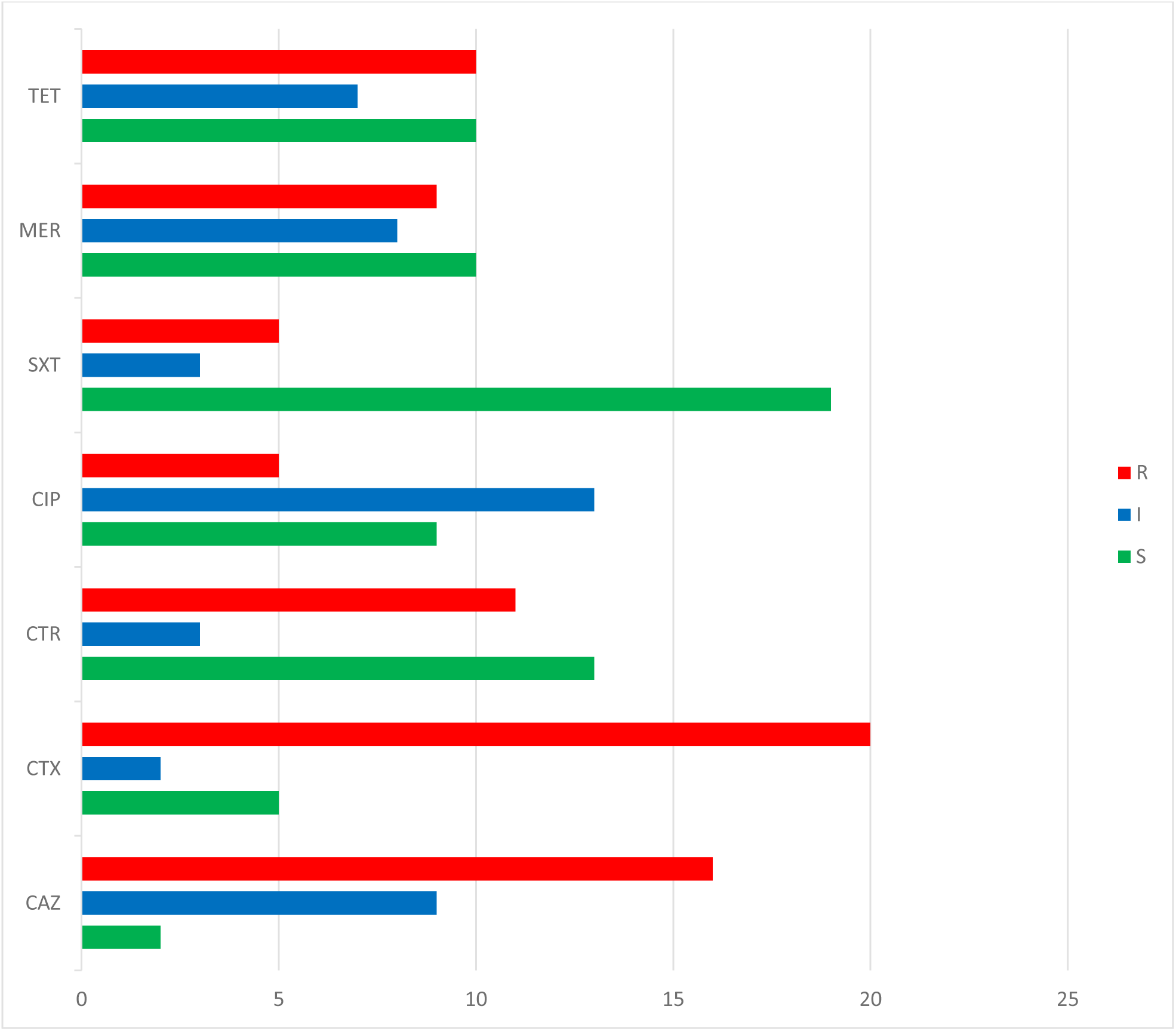
Antimicrobial susceptibility patterns of ESBL-PE isolated from vegetables sold in Aroge Gebeya, Hawassa, Ethiopia, 2024. **Note:** R: Resistance, I: Intermediate, S: Susceptible

**Table 5.** Antibiotics resistance patterns of bacterial isolates among vegetables in Aroge Gebeya, Hawassa, Ethiopia, 2024. The values are provided as numbers (n) and percentages (%).

| Isolate(n) | CIP | SXT | CAZ | CTX | CTR | FOX | GEN | MER | TET | ERY | CC |
| --- | --- | --- | --- | --- | --- | --- | --- | --- | --- | --- | --- |
| *S. aureus* (48) | 0(0.0) | 0(0.0) | NT | NT | NT | 14(29.2) | 0(0.0) | NT | NT | 2(4.2) | 7(14.6) |
| *Enterobacter spp.* (25) | 2 (8.0) | 8(32.0) | 14(56.0) | 11(44.0) | 6(24.0) | NT | NT | 4(16.0) | 7(28.0) | NT | NT |
| *Citrobacter spp.* (24) | 4(16.7) | 7(29.2) | 13(54.2) | 12(50.0) | 13(54.2) | NT | NT | 5(20.8) | 11(45.8) | NT | NT |
| *Escherichia coli* (17) | 3(17.6) | 1(5.9) | 5(29.4) | 9(52.9) | 2(11.8) | NT | NT | 2(11.8) | 5(29.4) | NT | NT |
| *Salmonella spp.* (16) | 3(18.8) | 3(18.8) | 8(50.0) | 7(43.8) | 8(50.0) | NT | NT | 8(50.0) | 4(25.0) | NT | NT |
| *Klebsiella spp.* (13) | 1(7.7) | 2(15.4) | 5(38.4) | 8(61.5) | 3(23.1) | NT | NT | 1(7.7) | 2(15.4) | NT | NT |
| *Providencia spp.* (9) | 2(22.2) | 2(22.2) | 3(33.3) | 6(66.7) | 4(44.4) | NT | NT | 4(44.4) | 1(11.1) | NT | NT |
| *Proteus spp.* (8) | 1(12.5) | 1(12.5) | 4(50.0) | 3(37.5) | 3(37.5) | NT | NT | 4(50.0) | 3(37.5) | NT | NT |
| Total (160) | 16(10.0) | 24(15.0) | 52(46.4) | 56(50.0) | 39(34.8) | 14(29.2) | 0(0.0) | 28(25.0) | 33(29.5) | 2(4.2) | 7(14.6) |

**Note:** CIP: Ciprofloxacin, GEN: Gentamicin, ERY: Erythromycin, TET: Tetracycline, FOX: Cefoxitin, CTR: Ceftriaxone, MER: Meropenem, CAZ: Ceftazidime, CTX: Cefotaxime, CC: Clindamycin, SXT: Trimethoprim-Sulfamethoxazole.

**Table 6.**
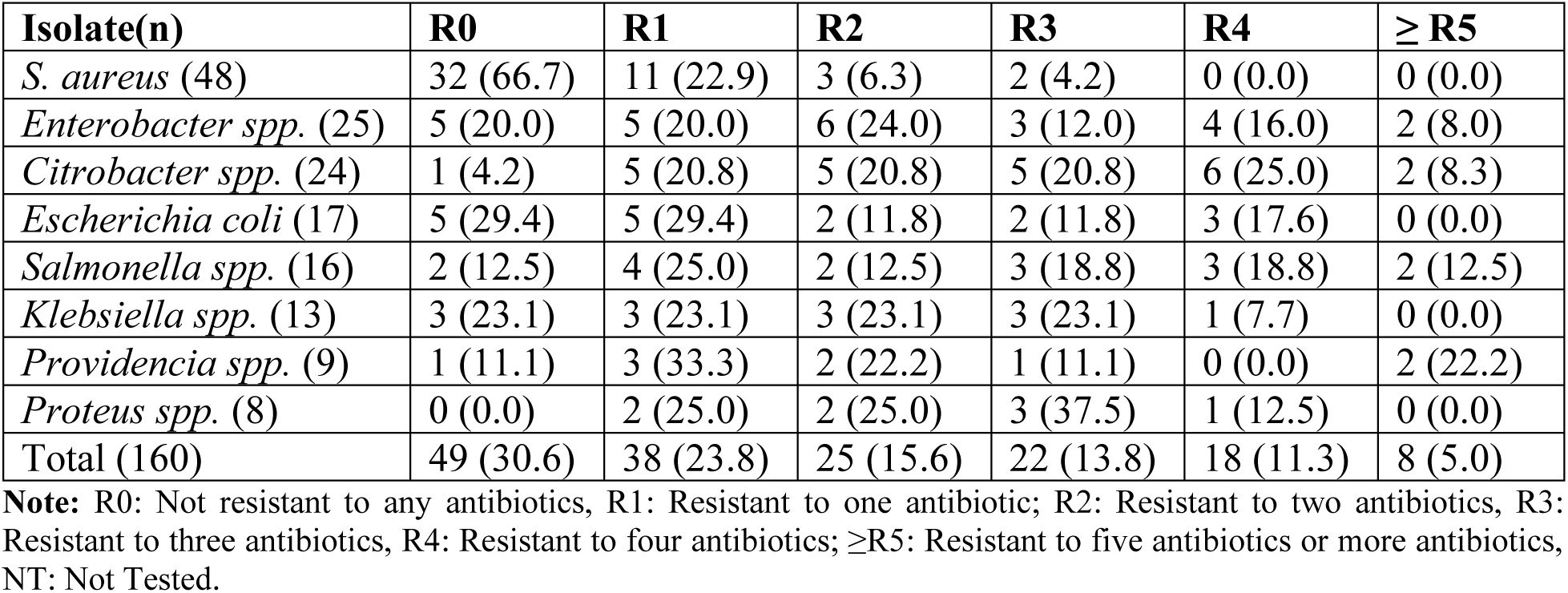
Multiple-drug resistance patterns of bacterial isolates among vegetables in Aroge Gebeya, Hawassa, Ethiopia, 2024. The values are provided as numbers (n) and percentage (%), n (%).

**Table 7.** Inducible clindamycin resistance observed among vegetables in Aroge Gebeya, Hawassa, Ethiopia, 2024. The values are provided as numbers (n) and percentage (%), n (%).

| *S. aureus* | ICR |
| --- | --- |
| MRSA (14) | 2 (14.3) |
| MSSA (34) | 0 (0.0) |

**Note:** ICR: Inducible Clindamycin Resistance, MRSA: Methicillin-resistant *S. aureus*, MSSA: Methicillin-susceptible *S. aureus*

### Factors associated with the overall prevalence of MRSA and/or ESBL-PE among raw vegetables

Vegetables sold by vendors who did not cut their nails were most likely to have a higher overall prevalence of MRSA and/or ESBL-PE (AOR = 4.123; 95% CI: 1.681 - 10.109, p = 0.002) compared to the vegetables sold by the vendors with trimmed nails (Table 8).

**Table 8.**
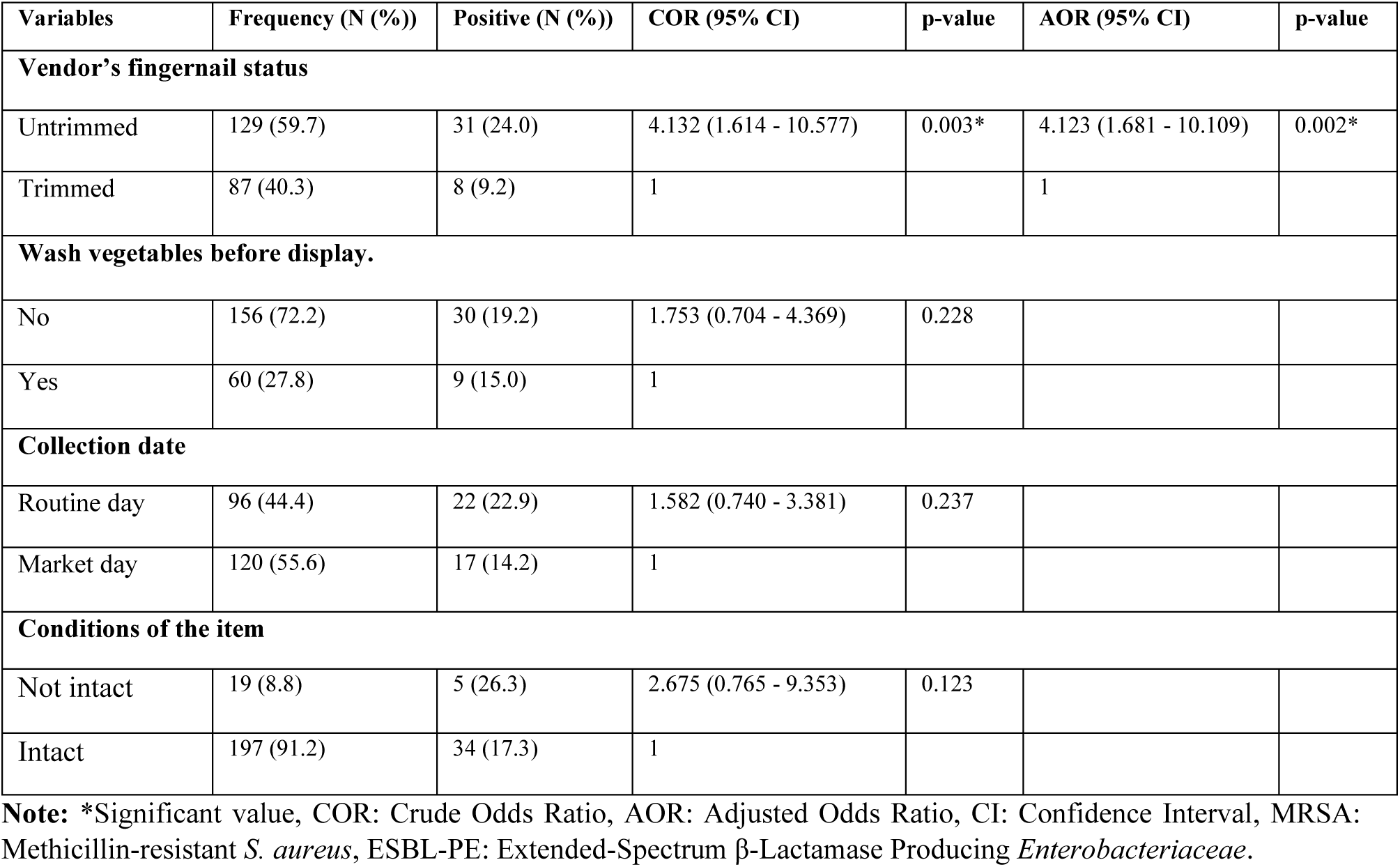
Factors associated with overall prevalence of MRSA and/or ESBL-PE among vegetables in Aroge Gebeya, Hawassa, Ethiopia, 2024. The values are provided as numbers (N) and percentages (%).

| Variables | Frequency (N (%)) | Positive (N (%)) | COR (95% CI) | p-value | AOR (95% CI) | p-value |
| --- | --- | --- | --- | --- | --- | --- |
| **Vendor's fingernail status** |  |  |  |  |  |  |
| Untrimmed | 129 (59.7) | 31 (24.0) | 4.132 (1.614 - 10.577) | 0.003* | 4.123 (1.681 - 10.109) | 0.002* |
| Trimmed | 87 (40.3) | 8 (9.2) | 1 |  | 1 |  |
| **Wash vegetables before display.** |  |  |  |  |  |  |
| No | 156 (72.2) | 30 (19.2) | 1.753 (0.704 - 4.369) | 0.228 |  |  |
| Yes | 60 (27.8) | 9 (15.0) | 1 |  |  |  |
| **Collection date** |  |  |  |  |  |  |
| Routine day | 96 (44.4) | 22 (22.9) | 1.582 (0.740 - 3.381) | 0.237 |  |  |
| Market day | 120 (55.6) | 17 (14.2) | 1 |  |  |  |
| **Conditions of the item** |  |  |  |  |  |  |
| Not intact | 19 (8.8) | 5 (26.3) | 2.675 (0.765 - 9.353) | 0.123 |  |  |
| Intact | 197 (91.2) | 34 (17.3) | 1 |  |  |  |

**Note:** \*Significant value, COR: Crude Odds Ratio, AOR: Adjusted Odds Ratio, CI: Confidence Interval, MRSA: Methicillin-resistant *S. aureus*, ESBL-PE: Extended-Spectrum $\beta$ -Lactamase Producing *Enterobacteriaceae*.

## Discussion

The presence of antimicrobial-resistant bacteria on raw vegetables is alarming the medical community, food industry, and public. This issue has escalated into a public health problem, with increasing food-borne illnesses in countries like Ethiopia. Despite their benefits, concerns about the safety and quality of vegetables are growing, highlighting gaps in our knowledge of pathogenic bacteria outside human and animal hosts (7,42). To our knowledge, there is limited information on the presence of MRSA and ESBL-PE in raw vegetables available in Ethiopian urban marketplaces.

This study found a high prevalence of *S. aureus* (30.0%; 48/160) among bacterial isolates in raw vegetables, indicating significant contamination. This finding suggests that raw vegetables can be a substantial source of *S. aureus* contamination, posing several public health risks. *S. aureus* contamination can lead to foodborne illnesses due to the production of enterotoxins, causing symptoms such as nausea, vomiting, diarrhea, and abdominal cramps. Similar studies conducted in Arba Minch and Debre Berhan, Ethiopia (7,9), as well as in Nigeria (43), Benin (44), Algeria (45) and Korea (46), reveal variations in prevalence rates. These differences can be attributed to factors such as variations in sampling methods, types of vegetables sampled, local hygiene practices, and agricultural methods. Understanding these differences is crucial for identifying the underlying causes and tailoring interventions to specific contexts. Vegetables can become contaminated with *S. aureus* through several pathways. Handling by farmers, vendors, and consumers can introduce *S. aureus* from the skin or respiratory tract. Contaminated soil, irrigation water, and the use of animal manure as fertilizer are also common sources of contamination (7).

In this study, 70.0% (112/160) were *Enterobacteriaceae* isolates including *Enterobacter spp*. 15.6% (25/160), *Citrobacter spp*. 15.0% (24/160), *E. coli* 10.6% (17/160), *Salmonella spp.* 10.0% (16/160), *Klebsiella spp*. 8.1% (13/160), *Providencia spp*. 5.6% (9/160) and *Proteus spp.* 5.0% (8/160) were isolated. Although the scope of the studies varied, other similar studies conducted in Ethiopia (7), Nigeria (47), Israel (29), Malaysia (48), Spain (49) and Romania (42) have also identified various enteric bacteria from raw vegetables. These differences could be due to the bacterial load in the environment, the culture techniques used, and the data collection period. Generally, the prevalence of enteric bacteria can be linked to coliform bacteria, which are typically found in feces and are common in areas with open defecation. Additionally, the use of human and animal manure as natural fertilizer by farmers contributes to the contamination of vegetables grown on these farms (7,47).

The study found the highest frequency of bacterial isolates in tomatoes (21.9%; 35/160), kale (18.1%; 29/160), and lettuce (16.9%; 27/160). This higher prevalence could be attributed to several factors, including the larger surface area and higher moisture content of these vegetables, which provide a conducive environment for bacterial growth. Additionally, these vegetables are often consumed raw, which means they are less likely to undergo processes that kill bacteria. These findings align with other studies that have shown leafy vegetables and tomatoes to be common sources of bacterial contamination (7,50).

In this study, the overall prevalence of MRSA and/or ESBL-PE among vegetables was 18.1% (39/216). Several factors might contribute to this prevalence, including poor hygiene practices during handling and processing and environmental contamination (47,48,51). To our knowledge, this is the first study to report the overall prevalence of MRSA and/or ESBL-PE in vegetables.

In this study, the prevalence of MRSA was 29.2% (14/48) among *S. aureus* isolates and 6.5% (14/216) among vegetable samples. This is lower compared to a study conducted in Debre Berhan, which found a prevalence of 36.4% (12/33) among *S. aureus* isolates and a nearly similar prevalence of 6.7% (12/180) among vegetable samples. This finding indicates the presence of *S. aureus* in Ethiopian vegetables, which harbor non-intrinsic resistance determinants, such as methicillin resistance (52). The vegetables analyzed in this study are typically consumed raw or slightly cooked, suggesting that consumers may be at risk of *S. aureus* infections.

The prevalence of ESBL-PE in this study was found to be 24.1% (27/112) among *Enterobacteriaceae* isolates and 12.5% (27/216) among vegetable samples, suggesting that raw vegetables could be a reservoir for resistance genes. This prevalence is higher than that reported in Debre Berhan, which was 18.5% (23/124) among *Enterobacteriaceae* isolates and a nearly similar prevalence of 12.8% (23/180) among vegetable samples (7). Recent research indicates that vegetables may act as potential conduits for the dissemination of resistance genes within the population. The overuse and unregulated use of beta-lactam antibiotics may have contributed to the emergence of ESBL producers.

In this study, 56.9% (91/160) of the pathogens were found to be MDR. Both *S. aureus* and *Enterobacteriaceae* exhibited a significant frequency of MDR. Similar findings have been reported in other investigations, with high MDR frequencies observed in Ethiopia (64.8%) (7), Nigeria (65.12%) (47), Romania (46.1%) (42) and Spain (33.5%) (53). These studies indicate that vegetables can act as vectors for a variety of MDR pathogens. The variation in bacterial-strain resistance rates across studies may be attributed to differences in geographical locations, the characteristics of fresh vegetable samples, and the selection of antibiotics and breakpoints.

This study reveals significant antibiotic resistance patterns among MRSA and ESBL-PE isolates in Ethiopia. Out of the total 14 MRSA isolates, 5 (35.7%) were resistant to clindamycin and 2 (14.3%) were resistant to erythromycin. Additionally, 2 out of 14 (14.3%) MRSA isolates showed ICR. Among the 27 ESBL-PE isolates, 20 (74.1%), 16 (59.3%), and 11 (40.7%) of ESBL-PE isolates were resistant to cefotaxime, ceftazidime, and ceftriaxone, respectively. The implications of these findings are significant for the treatment of infections, highlighting the need for alternative therapeutic options such as newer antibiotics or combination therapies.

Vegetables purchased from vendors with untrimmed fingernails exhibited a significantly higher overall prevalence of MRSA and/or ESBL-PE. Specifically, the odds of these vegetables having these bacteria were more than four times higher (AOR = 4.123) than those bought from vendors with trimmed nails. This finding was statistically significant (95% CI: 1.681 - 10.109, p = 0.002), highlighting a robust correlation. The implication here is profound. The condition of a vendor’s fingernails can be a key indicator of hygiene practices, and untrimmed nails can harbor harmful bacteria, which can easily transfer to the vegetables during handling. This emphasizes the critical role of vegetable handlers in maintaining hygiene standards to prevent cross-contamination and the spread of antibiotic-resistant bacteria. It underscores the importance of regular hand hygiene and proper nail care among vendors to protect public health.

### Limitations of the study

Molecular confirmation of the isolates was not performed, which limits our ability to analyze their genes for virulence and antibiotic resistance. Without molecular characterization, it is impossible to identify specific genetic markers or determine the clonal relatedness of MDR, MRSA, and ESBL-positive isolates. This gap restricts our understanding of the epidemiology and spread of these resistant strains.

Furthermore, the study did not compute common combinations of resistant antibiotics, which could have provided additional insights into resistance patterns and mechanisms. The unavailability of a sufficient number of antibiotic discs also limited the scope of antibiotic susceptibility testing, potentially affecting the accuracy and comprehensiveness of the resistance data.

These limitations might have affected the interpretation of the results. The absence of molecular data means we cannot confirm the genetic basis of resistance, which is crucial for tracking the spread of resistance and developing targeted interventions. Similarly, the lack of detailed resistance pattern analysis might obscure the true extent of multidrug resistance and the potential for cross-resistance among different antibiotics. Despite these limitations, the study provides valuable preliminary data on the prevalence and resistance patterns of bacterial isolates in raw vegetables, highlighting the need for further research with more comprehensive methodologies.

## Conclusion

This study revealed a high overall prevalence of MRSA and/or ESBL-PE in vegetables, with these bacteria showing strong resistance to commonly used antibiotics in Ethiopia. The increased antibiotic resistance among *S. aureus* isolates is particularly concerning for society. Notably, the isolation of MRSA from vegetables poses a public health risk for consumers. Additionally, many bacterial strains were resistant to multiple drugs and extended-spectrum β-lactamase. The findings highlighted poor handling and safety practices by vegetable vendors as potential risk factors for this high prevalence. These results suggest that raw produce, typically considered beneficial, can pose serious health risks.

### Recommendations

The findings of this study emphasize the importance of maintaining strict hygienic standards for raw vegetables in markets to reduce the overall prevalence of MRSA and/or ESBL-PE. This calls for the public health sector to ensure safe transportation, handling, and utilization of vegetables, along with continuous screening of market vegetables. The use of treated animal manure in agriculture also requires strict monitoring to prevent the emergence of antimicrobial-resistant bacteria in vegetables. It is crucial for the government to closely monitor both pre- and post-harvest activities of vegetable producers and sellers to minimize disease risks. Additionally, regular surveillance of antibiotic susceptibility is necessary to track changes in resistance patterns. Ensuring that regulated parties oversee vegetable safety and promoting safe handling and manufacturing practices throughout the vegetable production chain is essential. Future studies should focus on identifying the effects and occurrence rate of human colonization with antimicrobial resistance originating from raw vegetables to properly understand the true impact of the increasing prevalence of antibiotic-resistant genes along the food chain.

## Data Availability

All data produced in the present study are available upon reasonable request to the authors

## Abbreviations

AMR: Anti-Microbial Resistance
CLSI: Clinical and Laboratory Standards Institute
ESBL-PE: Extended-Spectrum Beta-Lactamase Producing *Enterobacteriaceae*
ICR: Inducible Clindamycin Resistance
MDR: Multi-Drug Resistance
MRSA: Methicillin-Resistant *Staphylococcus Aureus*
WHO: World Health Organization

## Acknowledgments

We would like to acknowledge the data collectors and laboratory staff working at the Microbiology Laboratory of HUCMHS. We also acknowledge all the study participants who consented to fill out the questionnaires and provided raw vegetable samples for laboratory analysis.

## Ethical approval and consent to participate

Ethical approval of the study was obtained from the institutional review board (IRB) of the College of Medicine and Health Sciences at Hawassa University (Ref. No: IRB/165/16). All participants were made aware of the study’s goal and each vegetable handler was provided with written informed consent. For minors, both consent and assent were obtained. All methods were performed in accordance with the relevant guidelines and regulations.

## Availability of data and materials

The data set used can be obtained from the corresponding author.

## Authors’ contributions

**DSA**: Proposal development, laboratory work, data analysis, drafting and final preparation of the manuscript.

**ABG**: Supervision of the study, data analysis and review of the manuscript.

**HSE**: Data collection, supervision of laboratory work and review of the manuscript.

**HTS**: Data collection, supervision of laboratory work and review of the manuscript.

**MDO**: Proposed the study, supervised the study, data analysis and review of the manuscript.

## Consent for publication

Not applicable

